# An Automated Entrustment Professional Activities Tracking System (EPA-TRAC) That Holistically Capture Competence: A Paradigm Shift to Programmatic Assessment

**DOI:** 10.1101/2022.06.05.22276005

**Authors:** Omayma Hamed

## Abstract

Competence is a complex phenomenon that requires comprehensive and holistic assessment. This could be only be achieved through a complete assessment program that navigates poles between assessment for learning, assessment of learning and assessment as learning. Assessment of competence is not an easy task; educationists resorted to unfolding competence into sub-sub-competences and assessing them as separate chunks which violated the purpose of competency-based education. This is attributed to the reduction of the rich information from an assessment to a numerical score. In addition, evaluations of chunks from different competency domains are collated together in a compensatory manner rather than in a conjunctive manner resulting in adding oranges to apples with resultant meaningless assessment information. A useful contribution to this whole issue is the introduction of entrustable professional activities (EPAs) which are considered portal for capturing and visualizing competences in a holistic manner. EPAs provide a better language to make education more task-based. This necessitated a different assessment paradigm “Programmatic Assessment”. This assessment paradigm ensures convergence of assessment data points that keep competence integrated into a whole meaningful picture. In this type of assessment model, the trainee has role in gathering, collating and reflecting on the various assessments in an informative way that triangulates evidence and translates them into learning goals. Hence longitudinal, information-rich feedback and reflection are crucial in such assessment system and could be collated in a developmental portfolio. In such model a variety of assessment tools could be used so long as they constitute a high utility index; assessment could be psychometric and workplace-based. Whether scores from psychometric assessments or enaction of feedback and learning goals are used for making judgment, remains the decision of the curriculum and assessment system designers according to context and purpose of assessment. Being complex to realize, an automated system (EPA-TRAC) is developed to capture that assessment system.

## 1 Introduction

The theoretical value of Entrustable Professional Activities (EPAs) assessment is rapidly expanding; however, an automated system is needed to facilitate its challenges. The aims of this study are to design, develop and examine an automated EPA-based assessment system for capturing the workplace performance of clinical trainees at the Armed Forces College of Medicine (AFCM). This research presents an evaluation for the proposed system to overcome the original knotty question in any EPA software activity for how to accommodate flexibility in practical EPA software training programs during any EPA-Competency-based program.

In 2005, Ten Cate introduced Entrustable professional activities (EPAs) to re-conceptualize performance in the workplace in a unifying manner across the continuum of medical training [1]. The EPAs activities have developed as a novel theoretical approach to evaluate the workplace performance in medical education [2]. It sizes upon three key concepts. The first one is professional activities in which the authentic units of work are to be carried out by a profession. The second is levels of supervision which is the level of support that a trainee needs to carry out a task in a safe and effective manner. The last one is the entrustment decisions in which the assignment of supervision levels to trainees to carry out a particular professional activity. There are several publications that trace in detail the breadth and depth of medical training programs that are EPA-based [3] [4] [5].

This research presents an EPAs tracking system to overcome general and local challenges. The general challenges include:

1. Collating back the assessment values of chunks from different competence domains in a meaningful way and linking them back to their generic competency domain is not impossible but requires a highly complex multi-dimensional mapping system.
2. Tracking each trainee along the learning trajectory to reach an entrustment decision is not an easy task.
3. Identifying whether trainees work on the supervisor’s feedback and learning goals and whether the supervisors’ feedback is effective in driving learning forwards is not an easy task.
4. To judge a trainee as entrusted to perform an EPA requires spotting a consistent performance at the expected level (minimum at three successive assessment points).
5. A regular reporting system is required to all stakeholders as per achievement of the required performance level of the professional activities (government-university administration-college administration-Competency Tracking Committee-Specialty Department Committee-Supervisors-Trainee).

Based on Root Cause Analysis (RCA) and Failure Mode & Effect Analysis (FMEA), the following risk points, concerning a new internship program, were prioritized according to their impact severity, occurrence, detection and control. The prioritized risk points and how they were overcome included:

1. The internship program is not assessed which makes it impossible to ensure competence of the graduating physicians *(Assessment System is required*.).
2. Large number of trainees makes it impossible to monitor and track the performance of each trainee *(Automated tracking system is needed*.)
3. Faculty illiteracy in using EPA-based observation cards and providing informative feedback *(Faculty development is crucial*.)
4. Departments resistance to change *(All stakeholders are included in a co-participatory manner in developing the training program)*
5. Generalization failure *(Piloting is a must*.)
6. Too much assessment tools *(The tools are reduced and simplified without jeopardizing their purpose*.)
7. Trainer-centred *(Workload minimized by allowing diverse sources to undergo assessment [supervisor-trainee-peer trainee])*.
8. Trainee not prepared *(Orientation & readiness month with the core competences in a simulated environment is set*.).
9. Lack of monitoring and auditing *(An automated tracking system is direly needed to realize the programmatic assessment system*.)

In this paper we focus on the development, piloting and evaluating an automated tracking system that facilitates the expert judgement on EPAs entrustment through visualization of performance whenever, and wherever needed.

## 2 EPA Tracking System Overview

This section will shade lights on how the proposed system works. The EPA tracking system works on professional’s opinions matter. The ultimate purpose of the two internship years program is aligned with the local and global functions required of a general practitioner. The proposed program aims at developing a general practitioner who is capable of accomplishing the required daily professional activities safely, competently and professionally. Hence, the approach to developing the program is anchored in pragmatic theory which emphasizes the validity of the outcomes by being more towards authenticity and real-life practice on the ground. Consequently, the program planners adopted the EPA-based approach to reconstruct the granular competence. The program used external and internal sources as benchmarks:

1. External Benchmark:
  1.1 Association of American Medical Colleges (AAMC): (13) Core EPAs [6].
  1.2 Royal College of Physicians & Surgeons in Canada (RC): (10) Transition to discipline & Foundation EPAs [7].
  1.3 General Medical Council Foundation Physician Capabilities [8]. All drives the curriculum development towards the EPA-based paradigm.
2. Internal Benchmark:
  2.1 National Academic Reference Standards [9].
  2.2 Central Egyptian medical Training Authority (CEMTA) guidelines for House Officers.

Both drives curriculum development towards the discrete competency-based paradigm.

In the automated system EPA-TRAC, the AAMC EPAs were mapped to the competency domains of the ACGME as well as to the competency domains of the other benchmarks. The system affords multidimensional mapping of the specific task to its relevant competency domain.

### 2.1 Automated EPA-TRAC Architecture

The system architecture shows how the system works, and interaction among its features. As shown in Figure 1, each intern is tracked through a digitized tracking system which is key for the success of programmatic assessment. Three inputs are fed into the tracking system:

**Figure 1.**
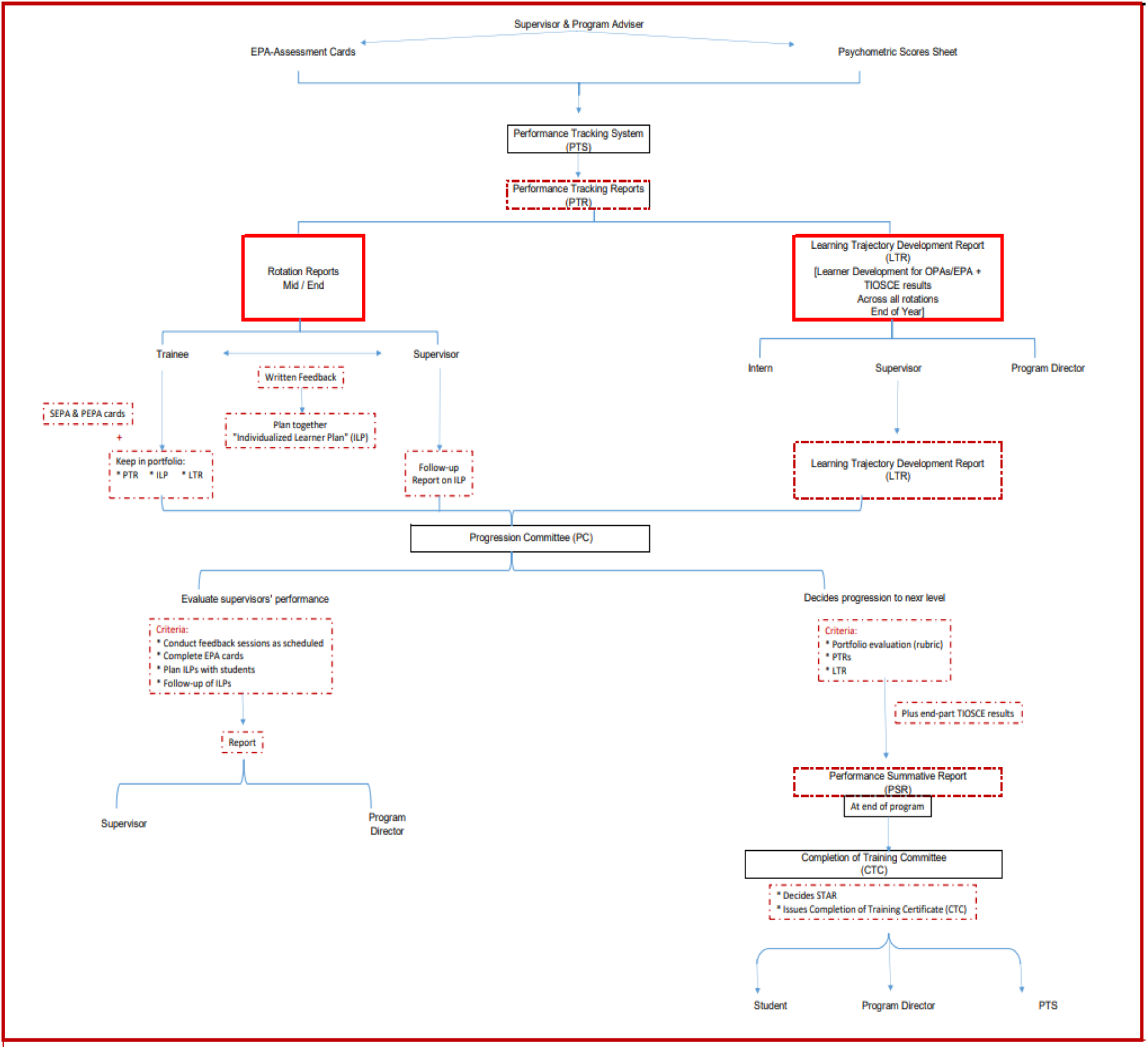
Automated EPA-Trac Architecture

- Interns’ scores in TIOSCE
- DEPA observation entrustment levels for each rotation
- Portfolio evaluation

The tracking system issues the following reports:

- To interns, supervisors, program advisers & Department Clinical Competency Committee (DCCC):
  - Mid-rotation report + Individualized Learner Plan (ILP)
  - End rotation report + Individualized Learner Plan (ILP)
- To Main Clinical Competency Committee (MCCC): (Colour coded)
  - Green final report for interns who are entrusted and succeeded their psychometric exams
  - Red final report for interns who are not entrusted or failed their psychometric assessments and have to repeat one or more rotations
  - The MCCC meets yearly and at the end of the program

## 3 Automated EPA-TRAC System Software

EPA’s Tracking user interface allows supervisor to complete their evaluations and submit them online. The system also features an auto-save function so that the evaluators may work directly in the application without worrying about losing their work. When the supervisor attempt to login to the system, his identity is checked by the login system as shown in Figure 2.

**Figure 2.**
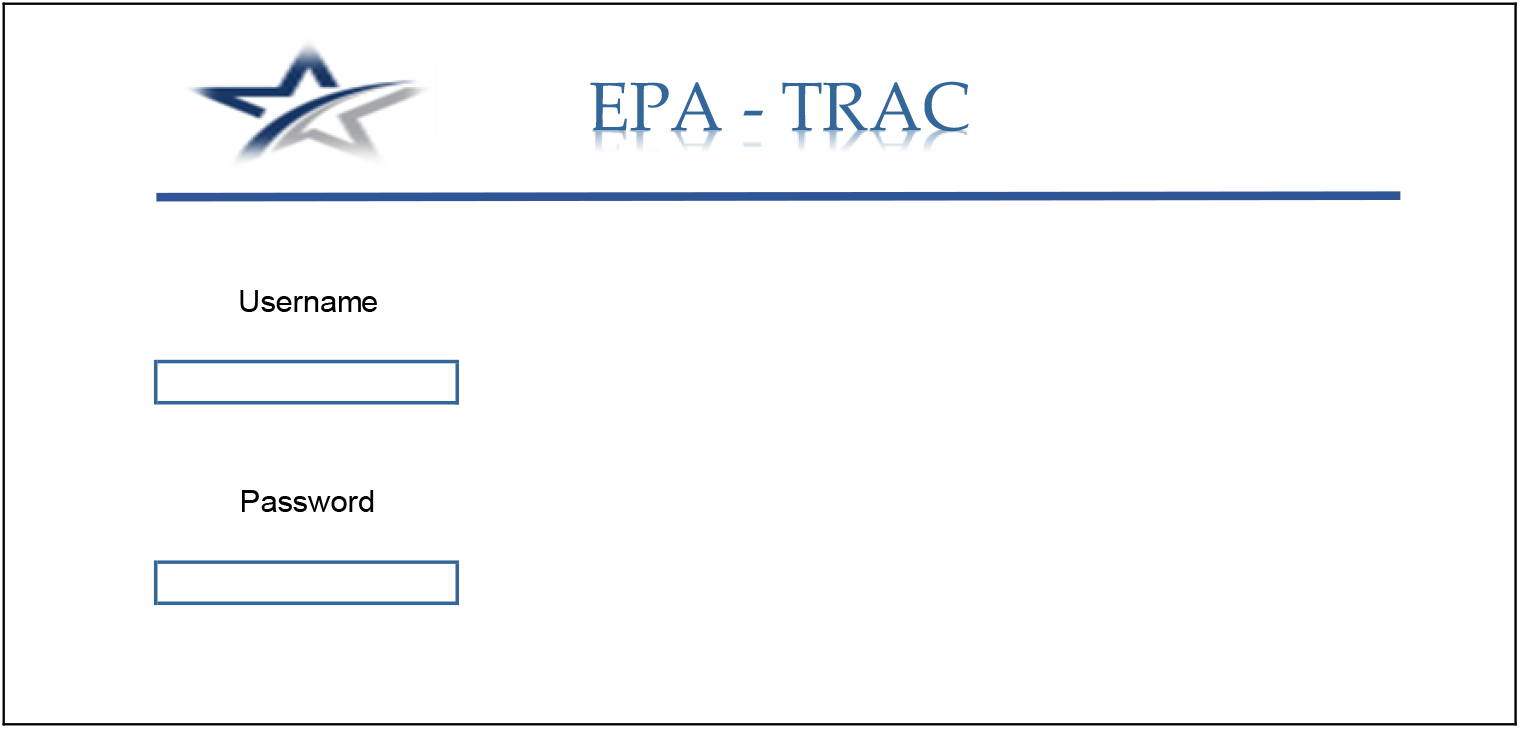
EPA-Trac login system

Figure 3, present the system dashboard. The dashboard presents a familiar user interface to manage trainee and supervisors assigned tasks, however, it displays all of the tasks in an organized manner. The dashboard has different functions for students, supervisors, and system admin. The top menu shows the number of active tasks. The whole context of the dashboard is changed based on the privileges of each user.

**Figure 3.**
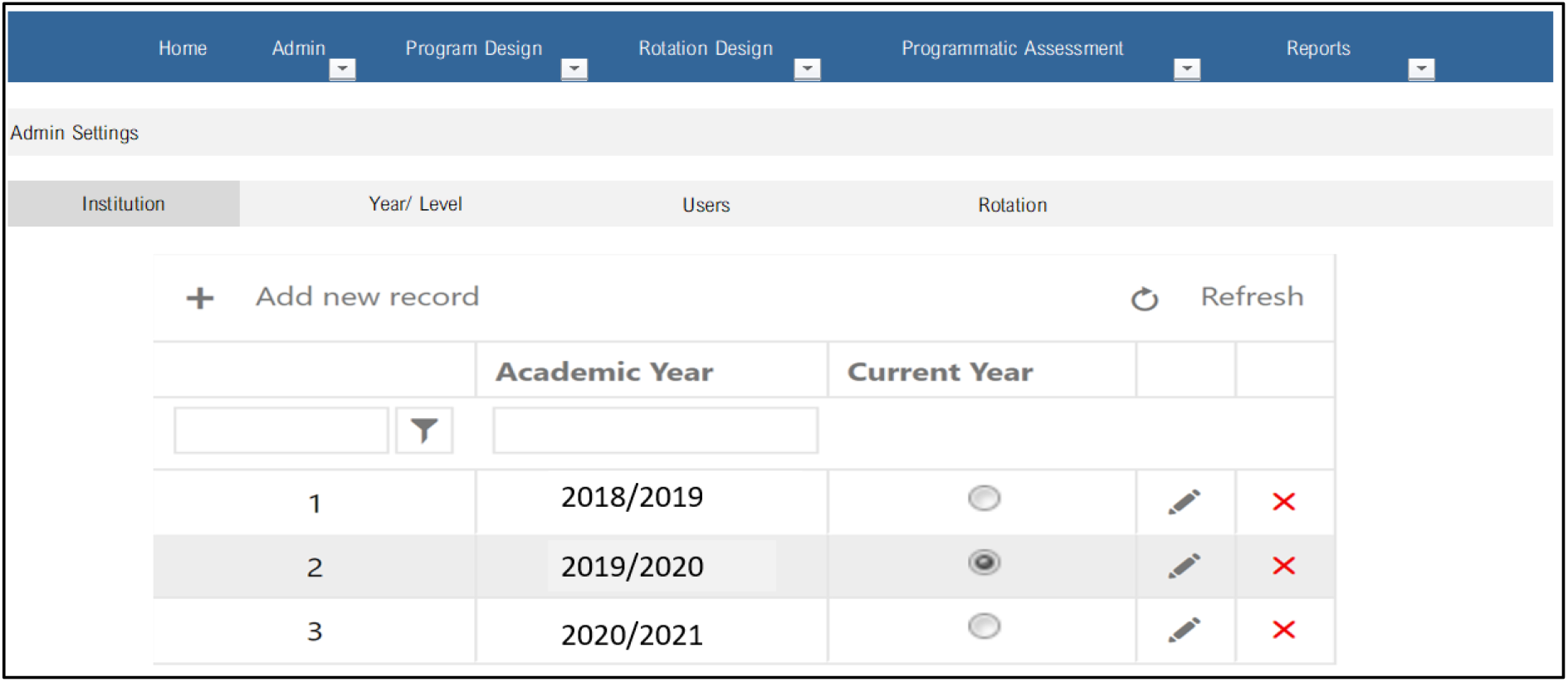
System Dashboard

For example, in the Internship Program Outline (IPO), Figure 3 shows the assessment Strategy & Reporting for the first year. Assessment is programmatic and consists of two arms:

A. **Psychometric assessment:**
  - At the end of the 10th week of each rotation, interns will sit for a Tasks Integrated Objective Structured Exam (TIOSCE) based on a blueprint that covers a representative sample of the tasks [Observable Practice Activities (OPAs)] in the EPAs selected by each specialty. TIOSCE overcomes the pitfall of compartmentalization of OSCE. It is an integrated approach to a case which mimics reality.
  - If the intern fails the exam, targeted remediation in the failed station is provided and the intern is allowed to re-sit the exam in the failed station at the end of the 11^th^ week. Another re-sit is provided at the end of the 12^th^ week.
  - If the intern fails the second re-sit, then he/she will repeat the rotation at the end of the 2nd year of the internship.
B. **Workplace-based assessment (WPBA):**
  - The intern (self-assessment) and peers (peer assessment) assess performance using Modified observation cards (SEPA & PEPA cards, respectively) for the daily activities that they experience. SEPA & PEPA cards are fed into the portfolio.
  - The preceptors and supervisors use Departmental EPA cards (DEPA cards) in scheduled mid-and end-rotation checkpoints as well as adhoc observation points. Feedback is narrative in the DEPA cards. Feedback is tagged with an individualized learner improvement plan (ILP) targeted towards the areas which require deliberate training.
  - Data from the psychometric assessment, portfolio SEPA, PEPA and DEPA cards are fed into the automated tracking system that issues a mid and end of rotation report for each intern as well as an annual report. This reduces the workload on the supervisors as well satisfies the philosophy of programmatic assessment.
  - The reports are sent to the intern, supervisor and program adviser. The report is discussed with the Department Clinical Competency Committee (DCCC).
  - Reports of interns with concerns will be delivered to the supervisor of the following rotation to keep an eye on those interns.
  - At the end of the program, a comprehensive annual report together with the trainee portfolio is delivered to the Progression Committee (PC) which issues a “Performance Summative Report” to the Completion of Training Committee (CTC) which decides a STAR (Statement of Awarded Reliability) according to Ten Cate and issues a Completion of Training Certificate.

**The system has more than one assessment tool such as:**

- SEPA Self-Assessment Cards
- PEPA Peer-Assessment Cards
- DEPA Observation Cards (Used for adhoc, mid and end of rotation)
- TIOSCE
  - Training of supervisors and preceptors, program advisers and interns on using the SEPA, PEPA & DEPA cards is mandatory.
  - Training of departments’ faculty responsible for TIOSCE in how to prepare and design TIOSCE stations and grade them.

## 4 How the EPA-TRAC Automated System works

The system offers four subjects for the supervisor to choose one of them: medicine, surgery, pediatrics, and Obgyn as shown in Figure 4. For example, if the pediatrics has been chosen, the system allows the supervisor to choose the EPAs and the corresponding OPAs as shown in Figure 5. During this session, the supervisor has to write his expectation level for each OPA and the system will calculate the average of the expected level for the whole EPA item.

**Figure 4:**
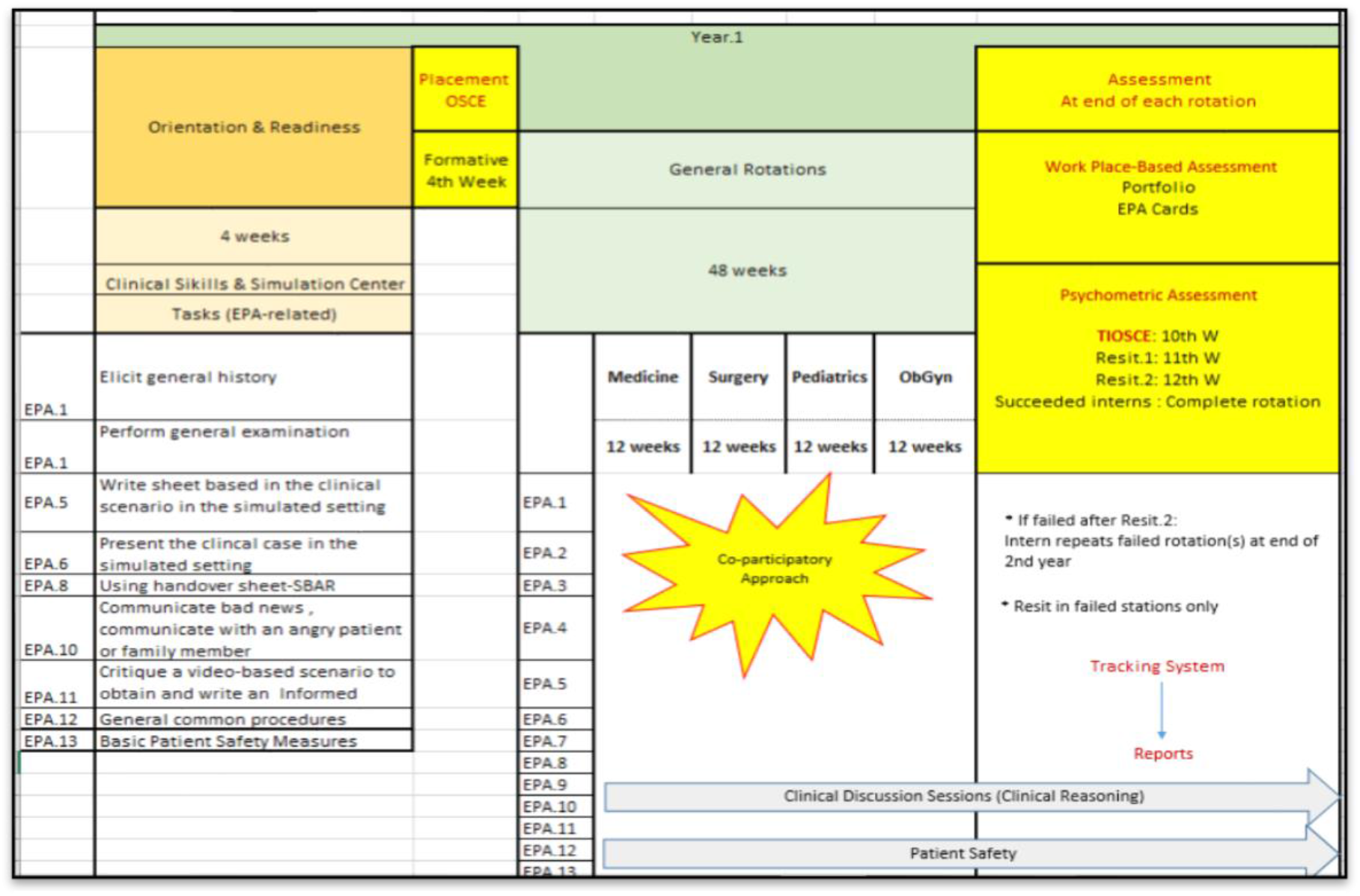
First year assessment Strategy

**Figure 5:**
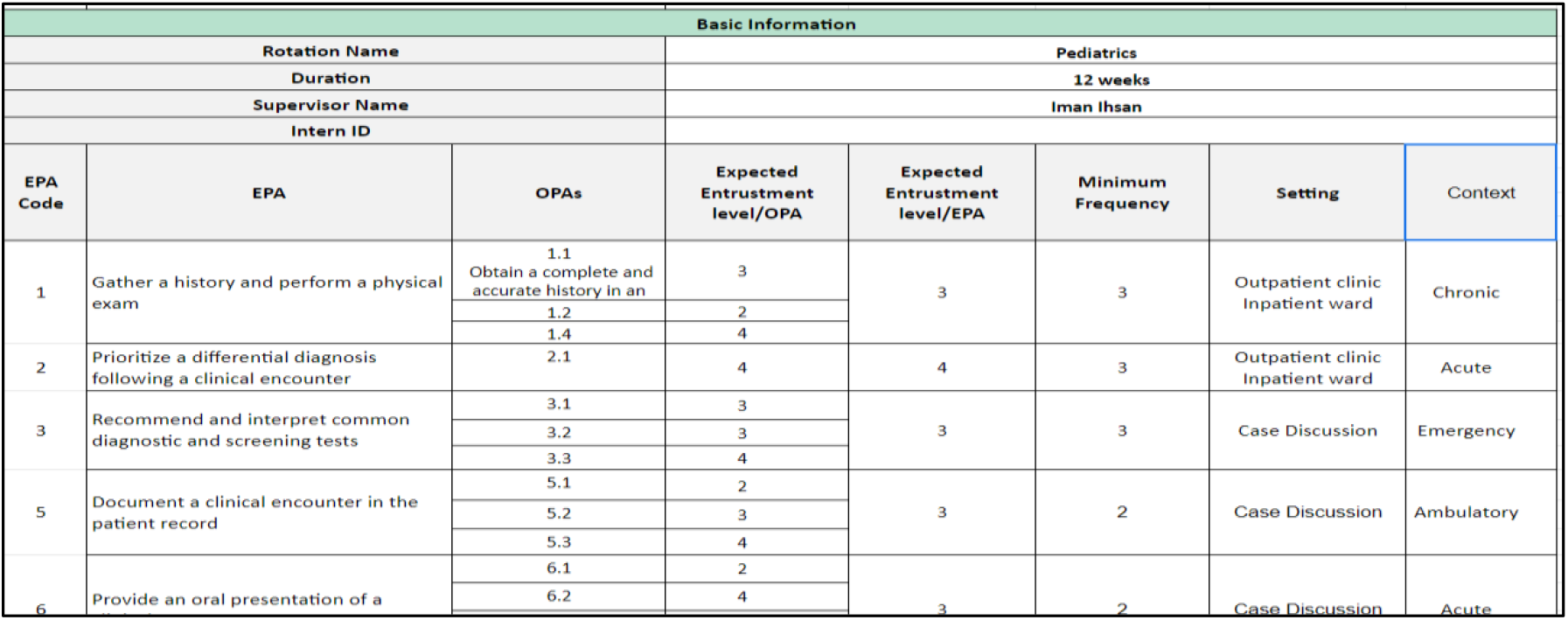
An excerpt from the Pediatrics Rotation designed using EPA-TRAC Automated System

During a duration of ten weeks, the supervisors use DEPA cards in scheduled mid- and end-rotation checkpoints as well as adhoc observation points. Feedback is narrative in the DEPA cards. Feedback is tagged with an individualized learner improvement plan (ILP) targeted towards the areas which require deliberate training. Figure 6, show how each rotation is divided into two equal durations and Card # (1) for the first half of the rotation displays weekly assessment points (week.1, week.2, etc) which are low stakes adhoc assessment points. The last week card is titled (Mid-rotation judgment card. This assessment point is a high-stakes one. Similarly, for the second half of the rotation

**Figure 6:**
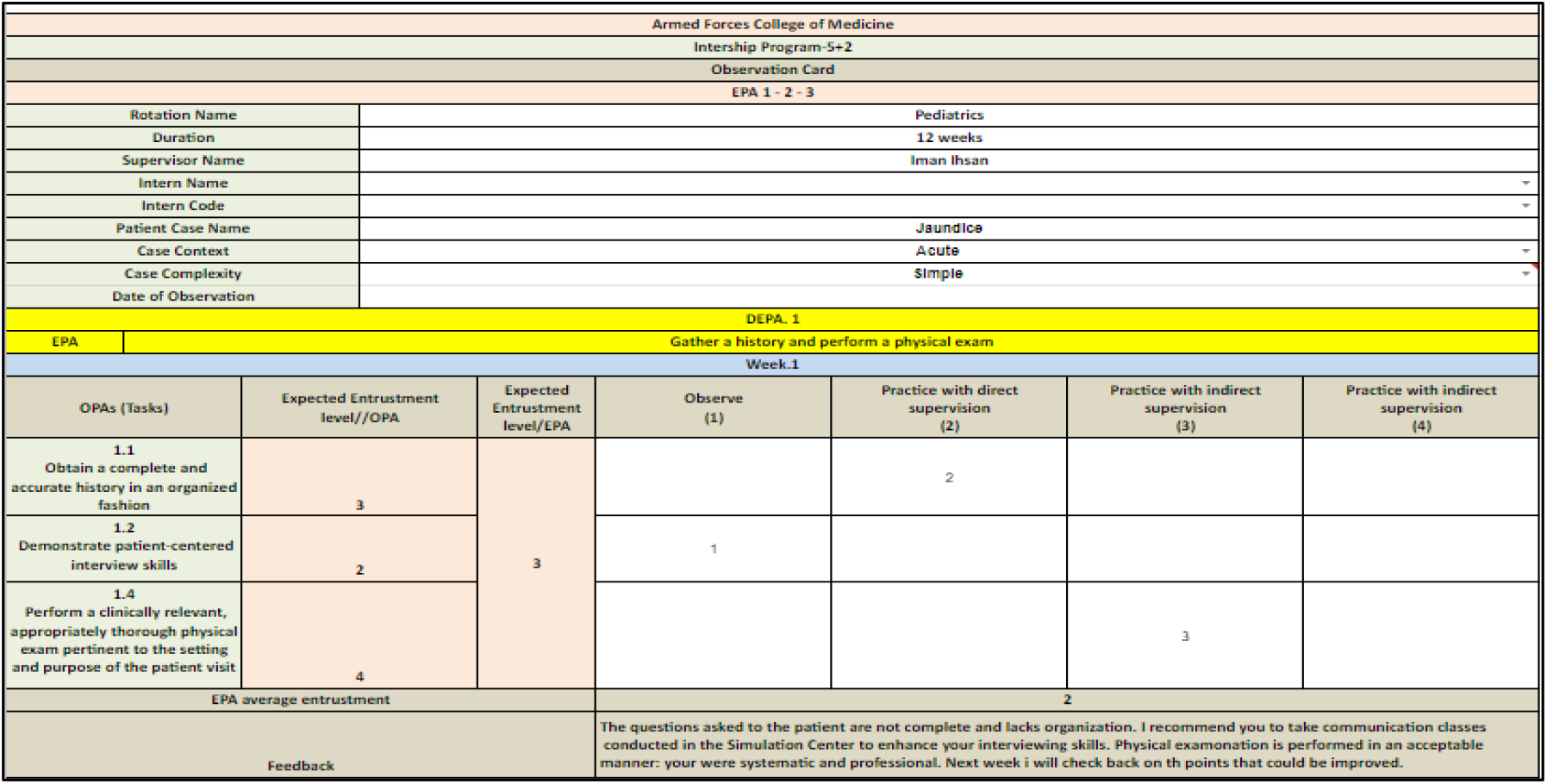
An excerpt from a DEPA Observation Card # (1) for EPAs 1-2-3 [Week.1]

The mid-rotation assessment is high stakes point as shown in Figure 7. The supervisor through the card can provide written narrative feedback and put an individualized learning plan (ILP) for each trainee. The supervisor also could raise any concerns regarding professionalism or patient safety and put an ILP with the trainee for it.

**Figure 7:**
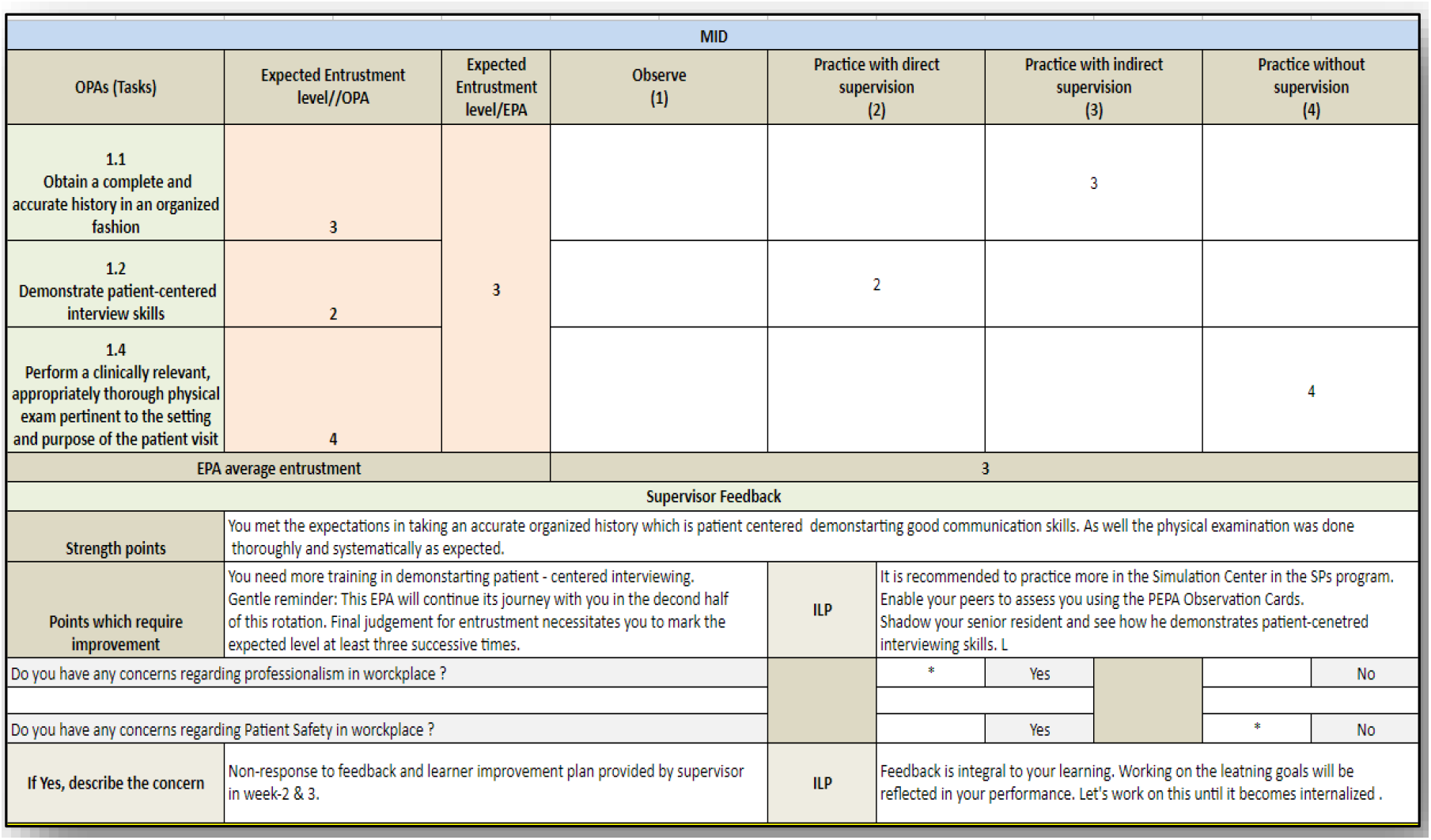
An excerpt from a DEPA Observation Card # (1) for EPAs 1-2-3 [Mid rotation]

Figure 8 shows an excerpt form for the mid-rotation report. This report displays the actual achievement of each EPA and its relevant OPAs benchmarked against the pre-determined expected level. It includes the supervisor’s feedback and Individualized Learner Plan (ILP) involved any professionalism or patient safety concerns are displayed in that report.

**Figure 8:**
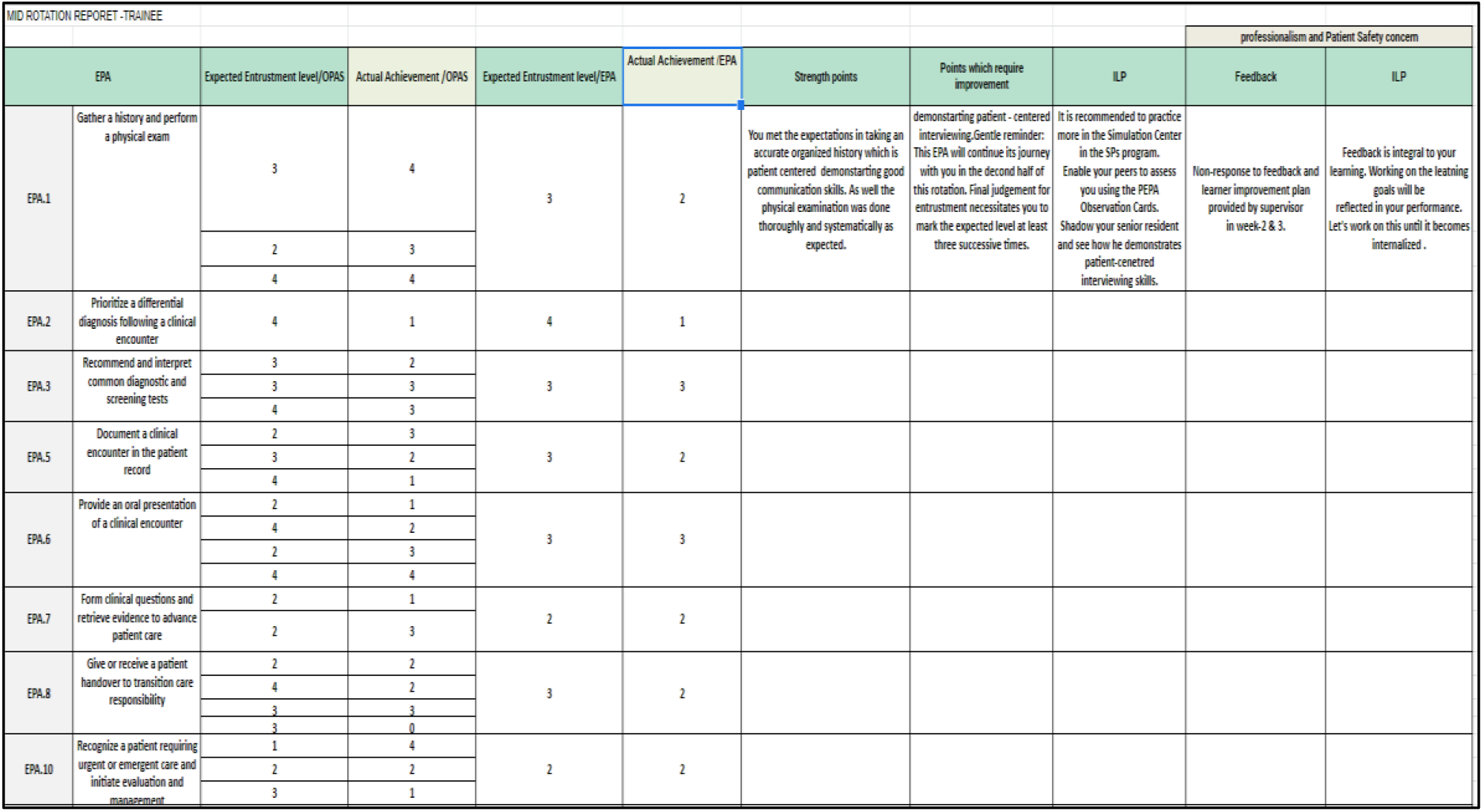
An excerpt from “Mid-Rotation Report”: [Trainee]

A similar report is issued at the end of rotation for each trainee to see their progress.

## 5 Experiment and results

The experiment is to design, develop and examine an automated EPA-based assessment system for capturing the workplace performance of clinical trainees at the Armed Forces College of Medicine (AFCM). Fifty trainees in the Pediatrics and ObGyn rotations were assessed in 2021/2022 academic year. For each of the fifty trainees (200) DEPA observation cards were used over 20 weeks and (6) performance reports were issued for both rotations. This makes a total of (10,000) cards and (300) reports for the (50) trainees in the pilot phase. This study is an initial study to produce and confirm a method for rating the characteristics of EPAs.

As shown in Figure 9, the developmental course of the actual achievement of each EPA for each trainee in all assessment points is displayed. The developmental course of the actual achievement of each OPA per EPA is also displayed.

**Figure 9:**
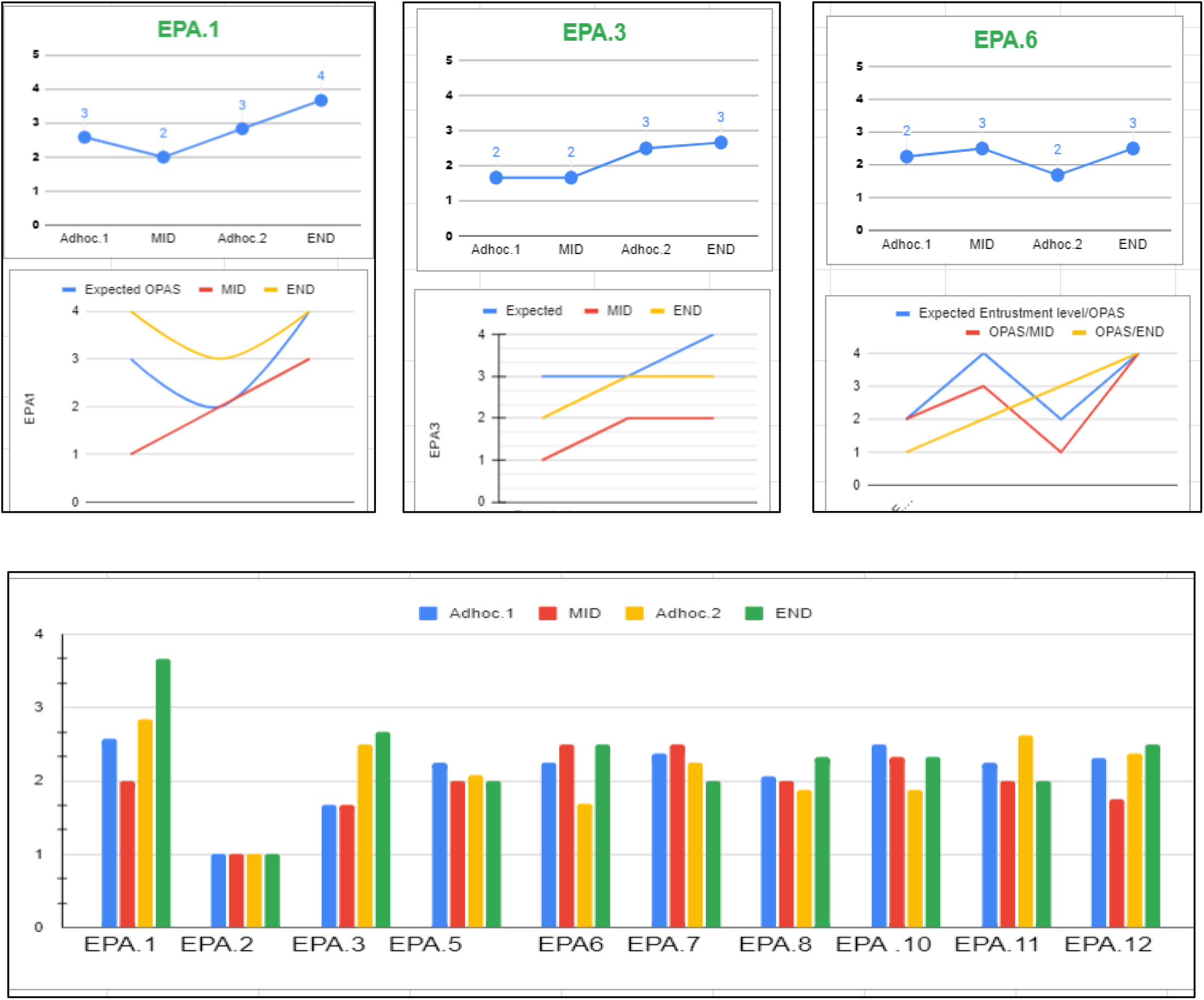
An excerpt from the “End Rotation report”: [Supervisor]

Figure 10 shows an excerpt from the TIOSCE scoring sheet. The TIOSCE exam is an academic assessment tool in the field of medical education. In this exam, User Satisfaction Survey to evaluate assessors’ attitudes is offered for each student. The student must pass all the stations. In case a student has failed in one or two station, a make-up station has to offer to him. In this sheet, the red coloured cells are the failed stations. Feedback with recommendations is written for each trainee and displayed at the annual report. Figure 11 presents the actual achievement at the end of each rotation of each EPA and its relevant OPAs is displayed with the feedback and ILP. The end of year TIOSCE results are also displayed with failed stations in red and recommended action before progression.

**Figure 10:**
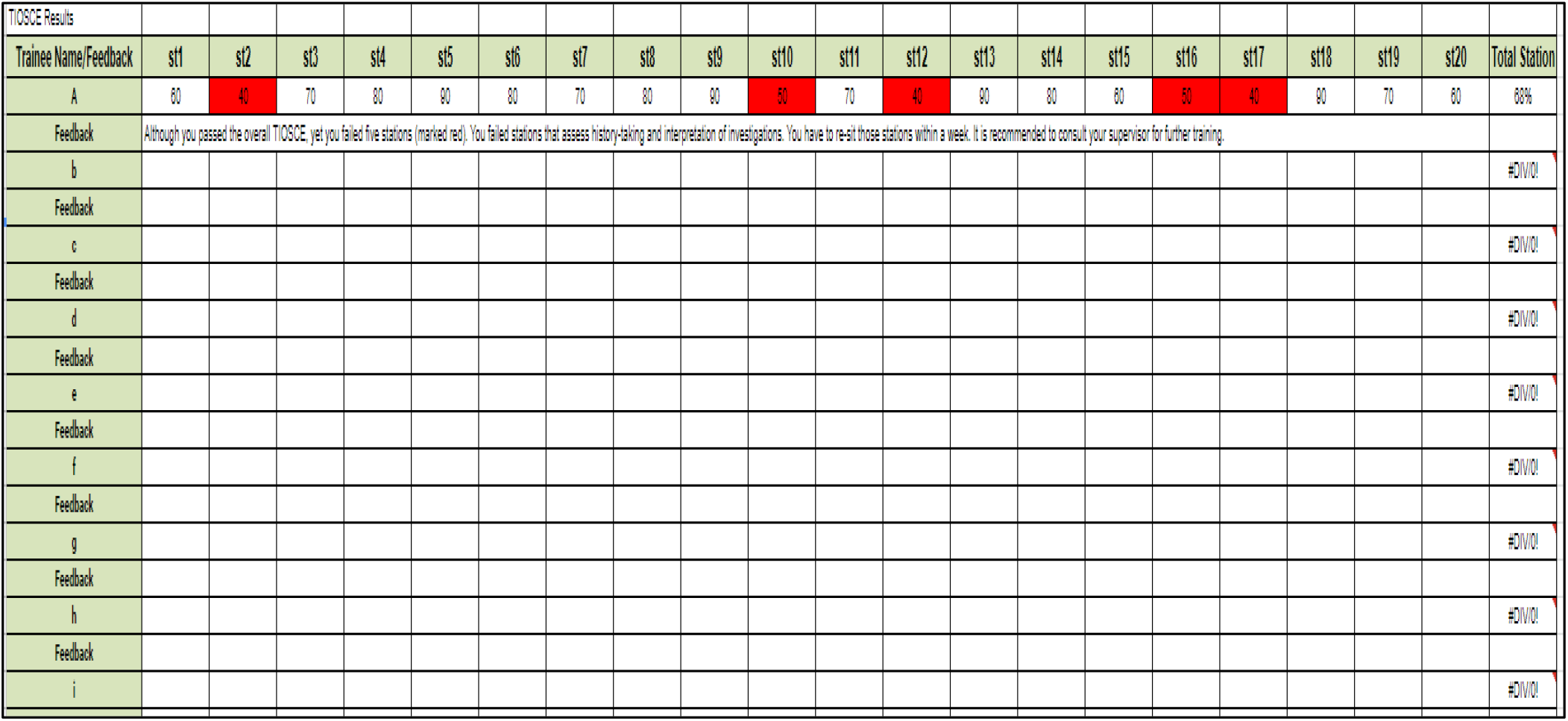
An excerpt from the TIOSCE scoring sheet

**Figure 11:**
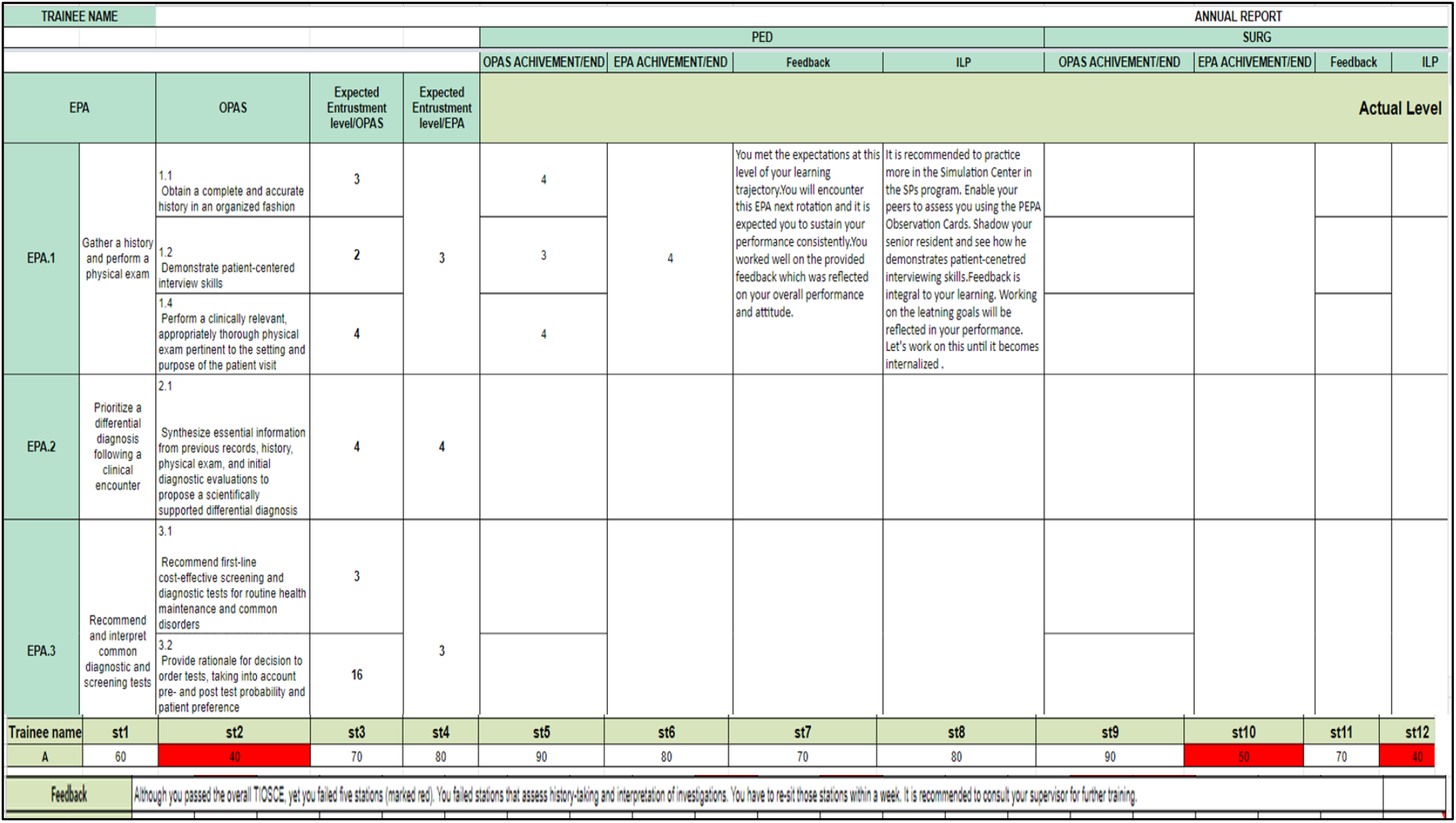
An excerpt from “Annual Report”

## Data Availability

All data produced in the present work are contained in the manuscript

## References

1. Ten Cate O. Entrustability of professional activities and competency-based training. Med Educ. 2005;39(12):1176–7.

2. Ten Cate O, Chen HC, Hoff RG, Peters H, Bok H, van der Schaaf M. Curriculum development for the workplace using Entrustable Professional Activities (EPAs): AMEE Guide No. 99. Med Teach. 2015;37(11):983–1002.

3. Holzhausen Y, Maaz A, Renz A, Peters H. Development of Entrustable Professional Activities for entry into residency at the Charité. GMS J Med Educ. 2019;36(1):Doc5.

4. Ten Cate O, Graafmans L, Posthumus I, Welink L, van Dijk M. The EPA-based Utrecht undergraduate clinical curriculum: Development and implementation. Med Teach. 2018:1–8.

5. Englander R, Flynn T, Call S, Carraccio C, Cleary L, Fulton TB, Garrity MJ, Lieberman SA, Lindeman B, Lypson ML, et al. Toward Defining the Foundation of the MD Degree: Core Entrustable Professional Activities for Entering Residency. Acad Med. 2016;91(10):1352–8.

6. Association of American Medical Colleges (AAMC), https://www.aamc.org/, (last accessed, May, 30, 2022).

7. Royal College of Physicians & Surgeons in Canada (RC), https://www.royalcollege.ca/, (last accessed, May, 30, 2022).

8. General Medical Council Foundation Physician Capabilities (GMCFPC), Practical skills and procedures, https://www.gmc-uk.org/-/media/documents/practicalskills-and-procedures-a4_pdf-78058950.pdf, (last accessed, May, 30, 2022).

